# *HYDIN* variants cause primary ciliary dyskinesia in the Finnish population

**DOI:** 10.1101/2024.05.28.24307879

**Authors:** Thomas Burgoyne, Mahmoud R Fassad, Rüdiger Schultz, Varpu Elenius, Jacqueline S Y Lim, Grace Freke, Ranjit Rai, Mai A Mohammed, Hannah M Mitchison, Anu I Sironen

## Abstract

Primary ciliary dyskinesia (PCD) is a rare genetic disorder characterized by chronic respiratory tract infections and in some cases laterality defects and infertility. The symptoms of PCD are caused by malfunction of motile cilia, hair-like organelles protruding out of the cell that are responsible for removal of mucus from the airways, organizing internal organ positioning during embryonic development and gamete transport. PCD is caused by mutations in genes coding for structural or assembly proteins of motile cilia. Thus far, mutations in over 50 genes have been identified and these variants explain around 70% of all known cases. Population specific genetics underlying PCD has been reported underlining the importance of characterizing gene variants in different populations for development of gene-based diagnostics and management. In this study, we identified disease causing genetic variants in the axonemal central pair component HYDIN. Three Finnish PCD patients carried homozygous loss-of-function variants and one patient had compound heterozygous variants within the *HYDIN* gene. The functional effect of the *HYDIN* variants was confirmed by immunofluorescence and electron tomography, which demonstrated defects in the axonemal central pair complex. All patients had clinical PCD symptoms including chronic wet cough and recurrent airway infections due to almost static airway cilia. Our results are consistent with the previously identified important role of HYDIN in the axonemal central pair complex and improve specific diagnostics of PCD in different ethnical backgrounds.

## Introduction

Primary ciliary dyskinesia (PCD) is an inherited disease with impairment of mucociliary clearance (MCC) leading to recurrent airway infections, bronchiectasis and in some cases severe lung damage in adult age. Mucus accumulation in the airways is caused by malfunction of motile cilia. Other syndromic features in PCD are situs inversus, otitis media, hearing loss, complex congenital heart disease, and more rarely hydrocephalus and retinitis pigmentosa (Shoemark and Harman 2021). Furthermore, PCD mutations may affect male and female fertility due to defective motile cilia in the oviduct and male efferent duct and cause defects in sperm tail development. Motile cilia line the airways and remove mucus and pathogens by a forward beating pattern, which is disrupted by mutations in genes regulating aspects of ciliogenesis or coding for dynein preassembly or structural proteins in motile cilia. Thus far variants in over 50 genes have been identified to cause PCD. The most commonly mutated genes code for components of the dynein arms, which are required for creating the movement of motile cilia (Fassad, Patel et al. 2020, Shoemark, Rubbo et al. 2021). The axonemal structure of motile cilia consists of 9+2 microtubules, inner and outer dynein arms, nexin links between the doublet microtubules, radial spokes connecting the outer microtubule doublets to the central pair and the central pair projection (Figure 1A).

**Figure 1.**
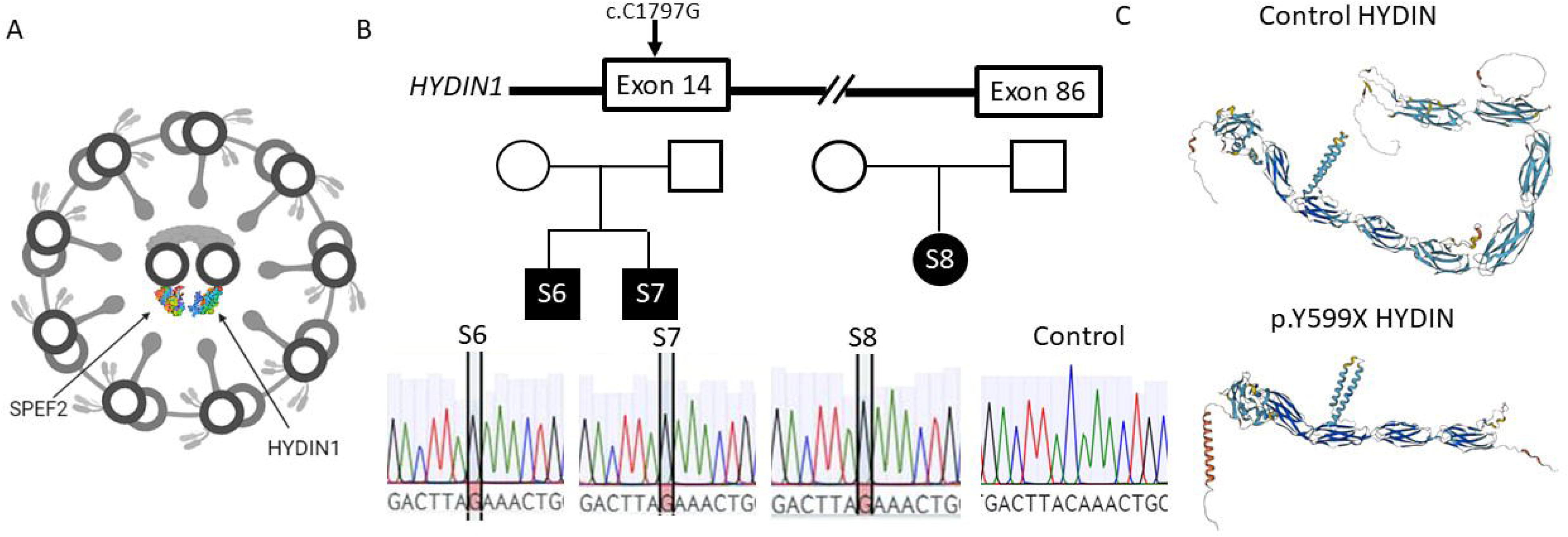
Stop gain variants in *HYDIN* cause PCD in Finnish patients. A. HYDIN and SPEF2 form a complex in axonemal central pair projection. B. Stop gain variant (c.1797C>G) in *HYDIN* exon 14 was identified in three Finnish PCD patients and confirmed by Sanger sequencing. C. The c.1797C>G variant results in truncated HYDIN protein product as visualized by the AlphaFold protein structure prediction tool (https://alphafold.ebi.ac.uk/).

The central pair projection is required for controlling the motility pattern created by the dynein arms and mutations in central pair complex genes such as *HYDIN* have been shown to result in stiff and static cilia (Olbrich, Schmidts et al. 2012). However, the central pair is not present in nodal cilia that govern left-right organ laterality and therefore mutations in genes coding for central pair proteins do not cause situs inversus (Best, Shoemark et al. 2019). Recently, it has been shown that lack of HYDIN causes depletion of SPEF2 from the ciliary central pair complex and thus immunostaining for SPEF2 can be used as a diagnostic tool for PCD patients with *HYDIN* mutations (Cindrić, Dougherty et al. 2020). Mutations in *SPEF2* mainly cause male infertility due to malformed and immotile sperm tails, but mild PCD symptoms have also been reported affecting the airways (Liu, Lv et al. 2019, Liu, Sha et al. 2019, Mori, Kido et al. 2022).

Two highly similar copies of *HYDIN* are present in the human genome, *HYDIN* which is transcribed from chromosome 16 (hg38 16:70,802,084-71,230,722) and a paralogue pseudogene *HYDIN2,* which is located on chromosome 1 (hg38 1:146,547,367-146,898,974). *HYDIN2* appears to be mainly expressed in the brain and fetal tissues with low expression in the lung, due to a neuronal promoter (Dougherty, Nuttle et al. 2017). *HYDIN2* is a duplicate of *HYDIN* exons 6-84 with short and long transcripts predicted to be transcribed, the most common isoform containing exons 6-19 (Dougherty, Nuttle et al. 2017). Due to their very high similarity, it is difficult to distinguish *HYDIN*-specific variants for diagnostics based on short read DNA sequencing. Therefore, RNA sequencing of nasal brushing biopsies can be potentially used to confirm causality of *HYDIN* variants in PCD patients. In this study we have identified novel and rare *HYDIN* variants in the Finnish population using exome and Sanger sequencing of DNA and RNA, and confirmed the functional effect of compound heterozygous variants using immunofluorescence and electron tomography.

## Methods

### Subjects

Blood samples were collected for DNA extraction from thirteen PCD patients, recruited at the University Hospitals of Turku, Kuopio and Tampere after written informed consent was given. Nasal brushing samples were collected for high-speed video microscopy analysis, immunohistochemistry and gene expression studies when possible. The study was ethically approved by the University of Turku Ethics Committee (ETMK 69–2017), London Bloomsbury Research Ethics Committee approved by the Health Research Authority (08/H0713/82), and the referring hospitals. The sample/patient IDs are not known to anyone outside the research group.

### Nasal nitric oxide analysis

For nasal nitric oxide (nNO) analysis, a CLD 88sp analyser equipped with a Denox 88 module for flow control was used (Eco Physics, Dürnten, Switzerland). If cooperativity was established, three consecutive trials were taken, from which the highest value was recorded. Nasal nitric oxide analysis was repeated on two different occasions.

### High-Speed Video Microscopy Analysis (HSVA)

Nasal epithelial cells were suspended in a DMEM medium and evaluated under a differential-interference microscope (Zeiss, Oberkochen, Germany) at x1000 magnification and cilia beat was recorded with a digital high-speed video (DHSV) camera (Hamamatsu Orca Flash 4.0, Hamamatsu Japan) with a frame rate of 256 Hz. The detailed protocol for HSVA can be found in Schultz et al. 2022. DHSV video sequences were played back frame by frame to determine the cilia beat frequency (CBF) by calculating the mean of all recorded cilia beat cycles. The cilia beating pattern (CBP) was determined by two independent expert operators. HSVA was repeated on two different occasions and averaged.

### Whole exome sequencing

The Nonacus Cell3 Exome panel was used for whole exome sequencing of patient DNA samples (https://nonacus.com/cell3tm-target). Unmapped reads were aligned to the current Human reference genome (GRCh38 build) by Burrows-Wheeler Aligner tool (Bwa-mem2 version) (Li and Durbin, 2010), SAM files were produced and indexed and converted to BAM format previous to marking and removing duplicates using Picard (https://broadinstitute.github.io/picard/). Subsequent analysis was executed following best practices guidelines for GATK from the Broad Institute (https://gatk.broadinstitute.org/hc/en-us). Firstly, base quality score recalibration (BQSR) was done to numerically correct individual base calls. Variant discovery was carried out in a two-step pipeline: variant calling with HaplotypeCaller followed by joint genotyping with GenotypeGVCFs. Once a multi-sample VCF file was obtained containing all definitive variant records, VariantRecalibrator was operated to fulfil Variant Quality Score Recalibration (VQSR) and refinement of the obtained variant callset.

Variants were individually selected for each sample. The called variants, including both single nucleotide variants (SNVs) and indels, were then annotated using ANNOVAR (http://www.openbioinformatics.org/annovar/). This tool enables functional annotation, and thus the final VCF files obtained contains detailed information for each variant site in the sample, such as their impact within a gene, predicted pathogenicity scores, minor allele frequency (MAF), zygosity status or reporting whether they have been recorded in large-scale databases like dbSNP.

### Variant prioritization

Variants were filtered for MAF <1%, their predicted functional impact on the encoded protein (missense, splicing, frameshift or nonsense) and frequency (<0.001) in the Genome Aggregation Database (gnomAD, broadinstitute.org). A list of genes with high expression during motile cilia development (n=652, reanalyzed gene list based on data in Marcet et al., 2011) was used to further filter variants in genes with potential roles in motile cilia. Finally, the pathogenicity of the identified variants was estimated with Combined Annotation Dependent Depletion tool (CADD, https://cadd.gs.washington.edu/snv) with CADD score >20 considered a significant pathogenicity score. Due to the known inheritance pattern of PCD homozygous variants, these were prioritized, but output files were also analyzed for the presence of compound heterozygous variants.

### RNA extraction and RT-PCR

RNA was isolated from nasal brushing samples using a Qiagen RNeasy kit (Qiagen) and RNA was reverse transcribed using a cDNA Reverse Transcription Kit (Applied Biosystems). cDNA was amplified using the TaqMan Fast Universal PCR Master Mix (Applied Biosystems). Primers used for RT-PCR are listed in Supplementary table 1.

### Sanger Sequencing

The identified variants were confirmed in the probands and available family members by Sanger sequencing. Primers flanking the mutation were designed using the NCBI Primer-BLAST tool (Supplementary table 1). The genomic or cDNA sequence was amplified using standard PCR conditions and predicted primer annealing temperature. The specificity of the PCR product was confirmed on agarose gel and purified using Exosap for Sanger sequencing.

### Transmission electron microscopy and electron tomography

A nasal brush biopsy was collected from a patient using 0.6-mm bronchial cytology brush. The sample was fix in 2.5% glutaraldehyde in cacodylate buffer before postfixing in 1% osmium tetroxide, and subsequently centrifuging in 2% agar to generate a pellet. Using increasing concentrations of ethanol followed by propylene oxide, the pellet was dehydrated before embedding in Araldite resin. 100 nm thick sections were cut and stained with 2% methanolic uranyl acetate and Reynolds lead citrate. Using a JEOL 1400+ transmission electron microscope (TEM) fitted with an AMT 16X CCD camera, conventional TEM images were acquired and 2D averaging was perform on TEM images using PCD-Detect (Shoemark, Pinto et al. 2020). To generate tomograms, perpendicular tilt series (dual axis) were collected using SerialEM (University of Colorado Boulder) with tilt increments of 1° over a range of ±60°. The tilt series were processed using IMOD (Kremer, Mastronarde and McIntosh 1996)to generate a dual axis tomogram.

### Immunofluoresence

Respiratory epithelial ciliated cells from nasal brushing of a PCD patient and a control sample were stained after blocking (10% BSA, in PBS) using DNAH5 (HPA037470, Cambridge Bioscience), RSPH4A (HPA031196, Sigma-Aldrich) and SPEF2 (HPA039606, Sigma-Aldrich) antibodies and colocalized with the cilia marker alpha-tubulin (322588, Invitrogen). After washes with PBST the slides were incubated with secondary antibodies Alexa Fluor 488 anti-mouse and Alexa Fluor 594 anti-rabbit (1:500). Slides were imaged using Zeiss Axio Observer 7 and deconvoluted using Huygens Deconvolution software (https://svi.nl/Huygens-Deconvolution).

## Results

### HYDIN variants identified in the Finnish population

Whole exome sequencing of Finnish PCD patients with chronic respiratory infections and otitis media (Table 1) identified one homozygous variant (NM_001270974:exon14:c.1797C>G:p.Tyr599*) in three patients and two heterozygous variants (NM_001270974:exon80:c.13801delG:p.Glu4601Argfs*17 and NM_001270974:exon76:c.12899G>C:p.Cys4300Ser) in one patient. These variants had a low frequency in the genomAD database and were predicted pathogenic using Polyphen, Sift and Provean, also having high CADD scores (Table 2). The homozygous variant was identified in two nonconsanguineous families and was confirmed using *HYDIN* specific primers for Sanger sequencing of patient DNA (Figure 1B). The variant results in a stop gain in exon 14 and a predicted truncated protein product, which is predicted to severely disrupt the HYDIN function. The protein 3D structure prediction tool AlphaFold (https://alphafold.ebi.ac.uk/) was used to demonstrate the effect of the stop gain variant on protein structure showing a clear reduction in size and a lack of several protein domains (Figure 1C).

**Table 1.** Patient characteristics of four patients with mutations in the *HYDIN* gene.

| Patient ID | Age | Sex | CBF Hz | Cilia beating pattern (HSVM) | Nasal NO | Clinical symptoms |
| --- | --- | --- | --- | --- | --- | --- |
| S10 | 20-25 | F | - | Unsynchronized, wavy | 15 | Recurrent sinusitis, otitis |
| S6 | 20-25 | M | 10.8 | Stiff, static | 29 | Recurrent sinusitis, otitis, respiratory infections |
| S7 | 20-25 | M | 0 | Stiff, static | 23 | Recurrent sinusitis, otitis, respiratory infections |
| S8 | 20-25 | F | 0 | Static | 5 | Recurrent sinusitis, otitis, respiratory infections |

**Table 2.** HYDIN variants identified in Finnish PCD patients.

| **Patient ID** | **Gene** | **Function** | **Variant** | **rsID** | **Zygozity** | **Frequency GenomAD** | **CADD Score** |
| --- | --- | --- | --- | --- | --- | --- | --- |
| S10 | HYDIN | frameshift deletion | HYDIN:NM_001270974:exon80: c.13801delG;p.E4601Rfs*17 | rs752405406 | het | 0.00002478 | 33 |
| S10 |  | nonsynonymous SNV | HYDIN:NM_001270974:exon80: c.G13800C;p.K4600N | rs751812896 | het | 0.00005122 | 16 |
| S10 | HYDIN | nonsynonymous SNV | HYDIN:NM_001270974:exon76: c.G12899C;p.C4300S | rs200169224 | het | 0.001577 | 23.8 |
| S10 | HYDIN | nonsynonymous SNV | HYDIN:NM_001270974:exon22: c.A3291G;p.I1097M | rs183427172 | het | 0.00367 | 5.758 |
| S6 | HYDIN | stop gain | HYDIN:NM_001270974:exon14: c.C1797G;p.Y599X | - | hom | - | 35 |
| S7 | HYDIN | stop gain | HYDIN:NM_001270974:exon14: c.C1797G;p.Y599X | - | hom | - | 35 |
| S8 | HYDIN | stop gain | HYDIN:NM_001270974:exon14: c.C1797G;p.Y599X | - | hom | - | 35 |

WES of patient S10 showed 4 variants within the *HYDIN* coding region of which two were predicted to be pathogenic (Table 2). A frameshift deletion (c.13801delG) was identified in exon 80 in cis with a nonsynonymous SNV (c.13800G>C). In trans with this allele, another nonsynonymous SNV was found in exon 76 (c.12899G>C), and was predicted to be damaging by Polyphen, Sift and Provean with a CADD score of 23.8 (Table 2). An additional benign nonsynonymous SNV was detected in exon 22 (c.3291A>G).

### HYDIN variants confirmed by Sanger sequencing and RNA expression

The stop gain variants were confirmed by Sanger sequencing of patients DNA using *HYDIN* specific primers (Supplementary table 1, Figure 1B). The similarity of *HYDIN* and *HYDIN2* across exons 72 to 85 hindered the confirmation of the identified variants in patient S10 since no *HYDIN* specific DNA primers could be designed. Therefore, we extracted RNA from the patient and her mother for transcript sequencing to confirm the variants for which we failed to find *HYDIN* specific primers for DNA sequencing. Sequencing of the variants at RNA level confirmed the presence and heterozygosity of both potentially causative *HYDIN* variants in the patient (Figure 2A) and the mother was a carrier of variant c.12899G>C. Sanger sequencing also showed a variant c.12900C>T predicted to result in a nonsynonymous SNV (Figure 2A), which did not affect the protein sequence. Therefore, we conclude that the PCD in patient S10 may likely be caused by variants c.13801delG and c.12899G>C. Furthermore, *HYDIN* expression across exons 76-81 and in the end of the gene exons 85-86 was dramatically decreased compared to the control (Figure 2B).

**Figure 2.**
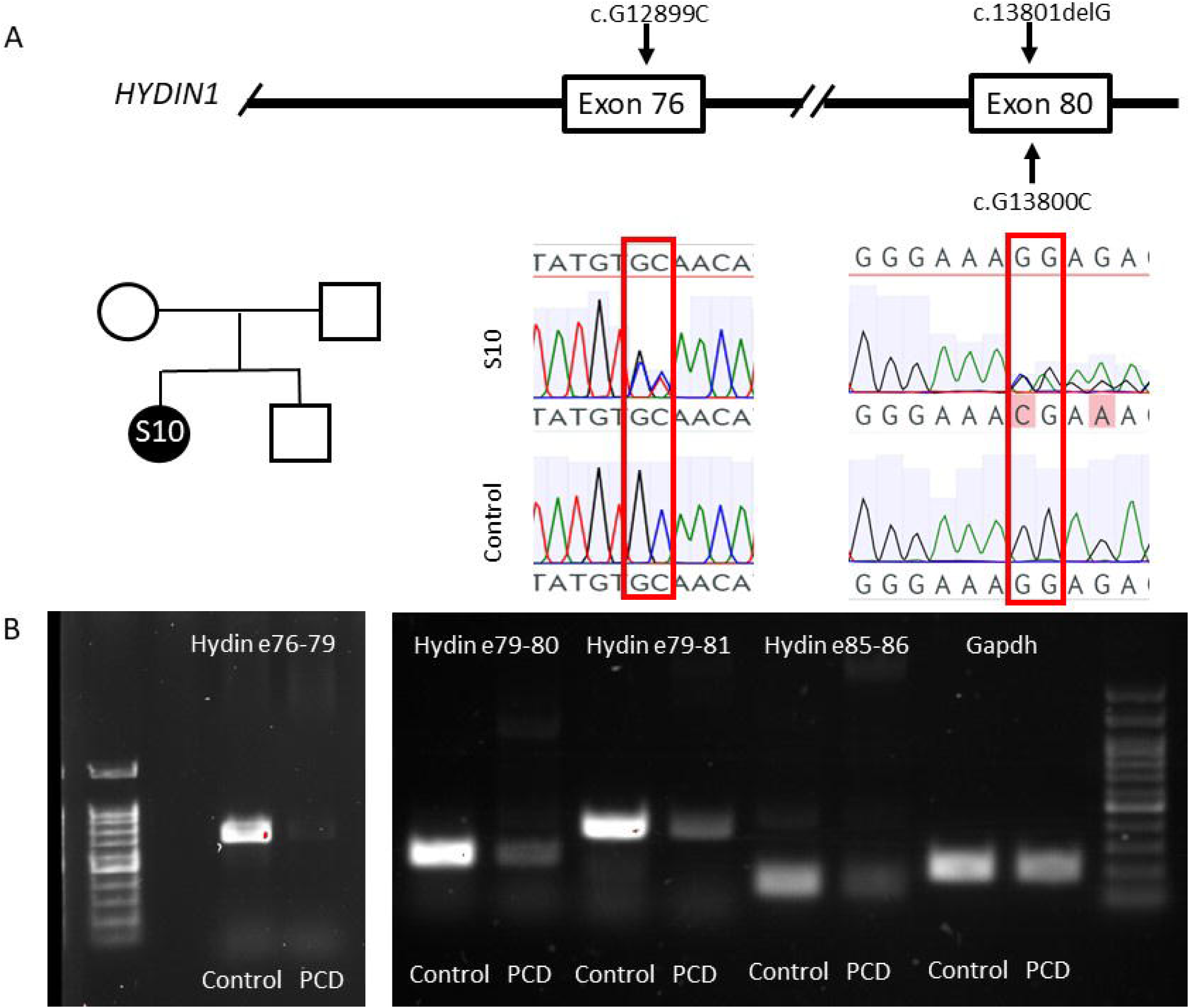
Compound heterozygous variants result in decreased *HYDIN* expression in a PCD patient. A. Sanger sequencing of patient RNA confirmed heterozygosity of the identified pathogenic *HYDIN* variants. B. HYDIN expression was decreased in the patient with HYDIN variants c.12899G>C and c.13801delG compared to control. *GAPDH* was used as a loading control.

### The axonemal central pair structure is affected in a patient with compound heterozygous HYDIN variants

To further validate the causality of the heterozygous *HYDIN* variants we investigated the structural defects in the ciliary axoneme of the patient’s nasal sample. TEM showed an apparently normal microtubular organization in the ciliary axonemes and the presence of dynein arms (Figure 3A), which was also confirmed by positive immunostaining with outer dynein arm component DNAH5 (Figure 4A). Furthermore, the radial spokes appeared unaffected as demonstrated by the presence of the radial spoke component RSPH4A (Figure 4A). Electron tomography has been demonstrated as a useful tool for identification of central pair structural defects (Shoemark, Burgoyne et al. 2018) and thus we utilized this method to confirm the expected disruption of the central pair complex arising from *HYDIN* mutations. Tomograms showed a lack of the proximal projection of the central pair complex in the patient with compound heterozygous variants p.Glu4601Argfs*17 and p.Cys4300Ser (Figure 3C), which was not obvious by the conventional TEM (Figure 3A). PCD-Detect (Shoemark, Pinto et al. 2020) was also able to identify the central pair defect in the patient compared to control (Figure 3D) demonstrating the usefulness of this tool in diagnostics. Since it has been previously shown that HYDIN is required for localization of SPEF2 in the axonemal central pair projection (Cindrić, Dougherty et al. 2020) we used SPEF2 immunostaining to confirm the lack of the HYDIN/SPEF2 protein complex in the axonemal central pair. SPEF2 was localized predominantly of cilia in airway epithelial cells from a healthy control individual, but was completely depleted in patient’s cilia (Figure 3B).

**Figure 3.**
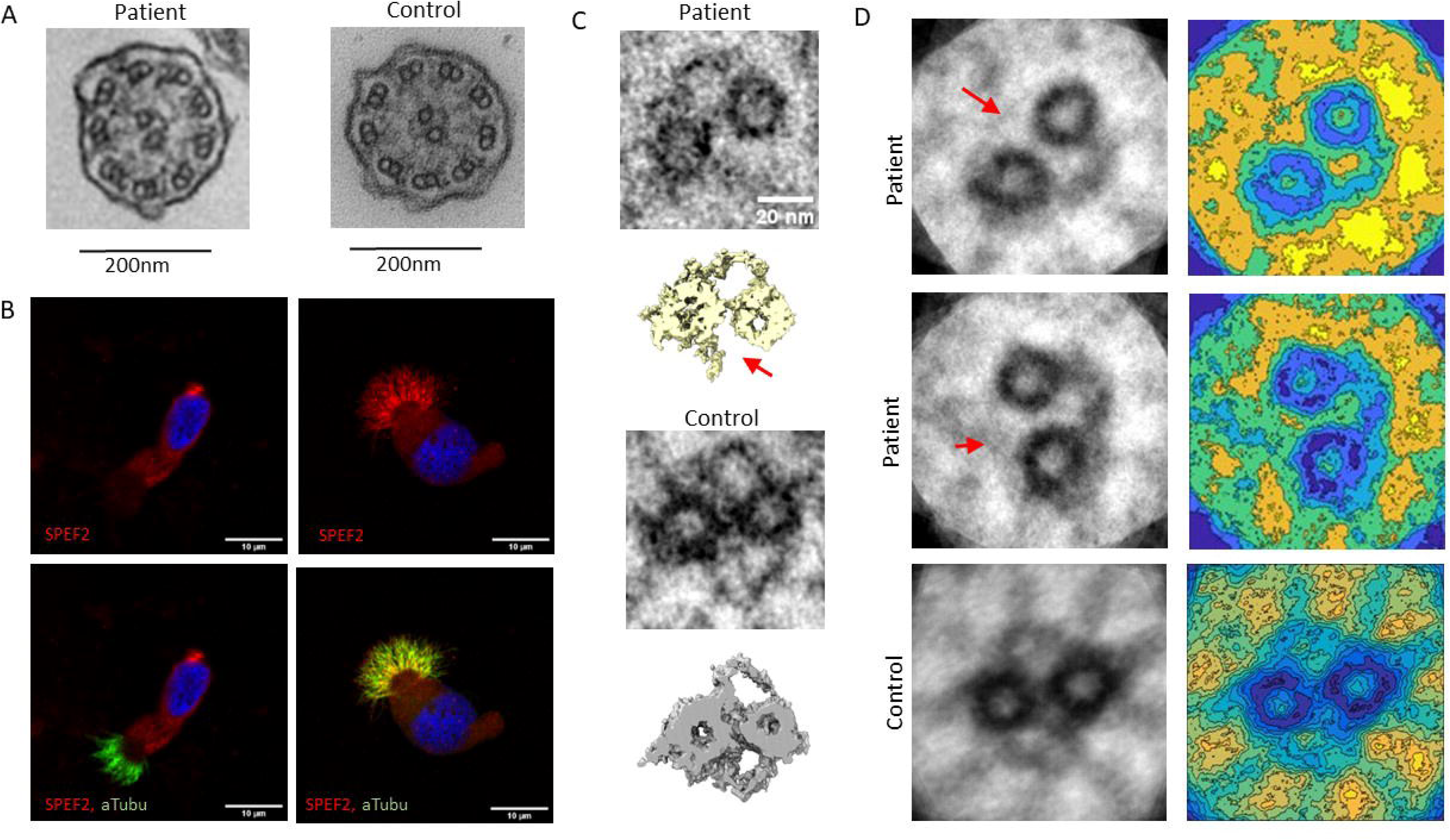
The axonemal central pair projection is affected in the PCD patient with compound heterozygous *HYDIN* variants. A. TEM showed apparently normal microtubular structure and the presence of dynein arms and radial spokes in the patient’s axonemal cross sections. B. SPEF2 staining was depleted in cilia of the patient’s nasal brushing samples. C. Electron tomography showed a lack of the SPEF2/HYDIN complex in the patient’s central pair projection. D. 2D image averaging using PCD Detect identified the lack of the SPEF2/HYDIN complex in the patient’s central pair.

**Figure 4.**
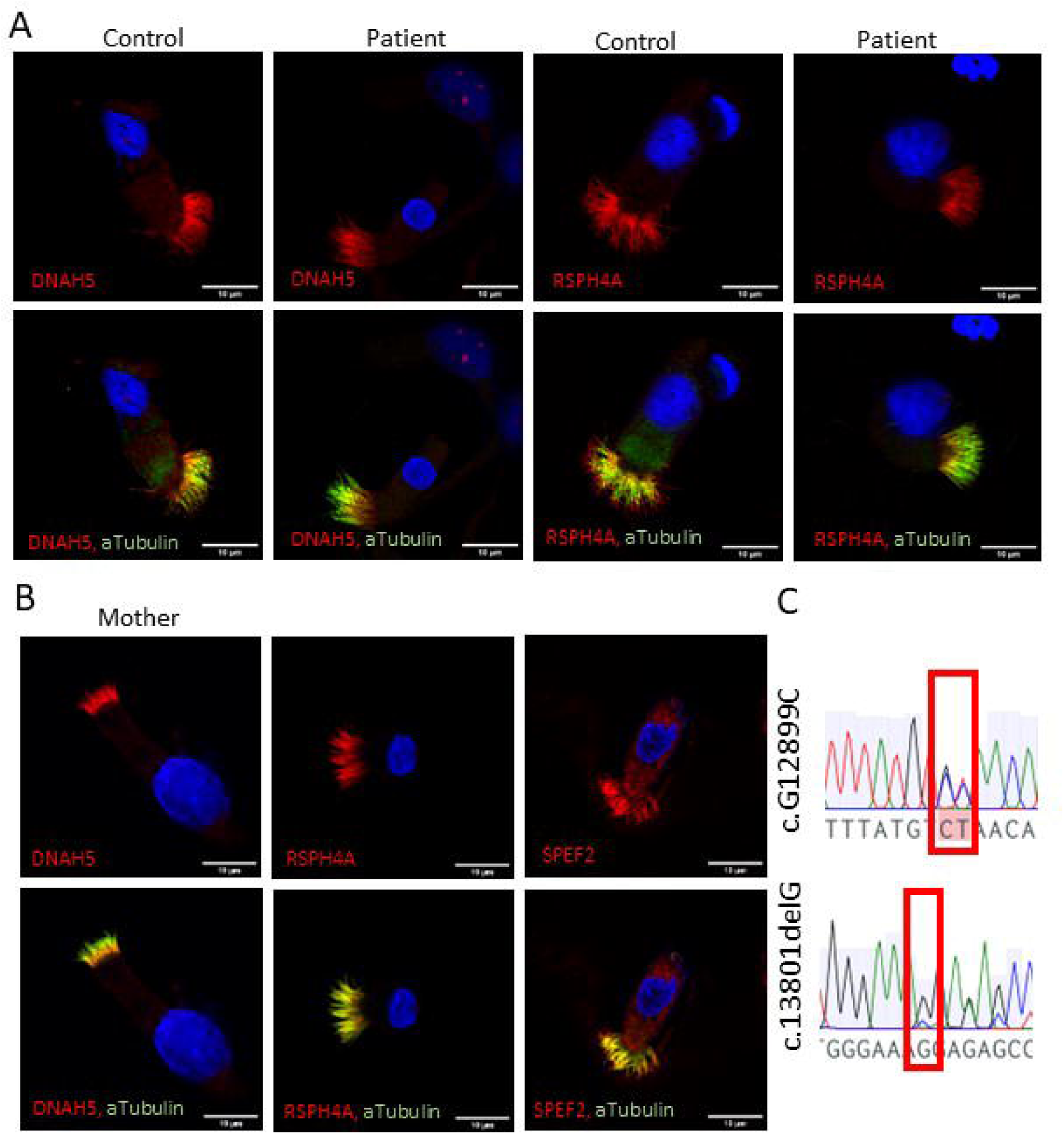
Immunofluoresence confirmed presence of the axonemal structures and Sanger sequencing the inheritance pattern of one *HYDIN* variant. A. Outer dynein arm protein DNAH5 and radial spoke protein RSPH4A protein localization in PCD patient with c.12899G>C and c.13801delG variants was comparable to the control. B. SPEF2, DNAH5 and RSPH4A were present in the airway cilia of patient’s mother. C. Sanger sequencing confirmed the inheritance pattern of the variant c.12899G>C from the mother.

Furthermore, we screened for the presence of the identified causative variants and protein localizations in the nasal sample of the mother of the patient. Localizations of SPEF2, DNAH5 and RSPH4A were comparable to a control sample (Figure 4B). Sequencing of the *HYDIN* variants showed inheritance of the c.12899G>C variant from the mother (Figure 4C). These results strongly support the conclusion that the identified *HYDIN* variants in patient S10 are causative for PCD and confirm the effect of the variants on HYDIN localization to the ciliary axoneme and the lack of functionality of the protein product.

### HYDIN mutations cause stiff cilia motility in Finnish PCD patients

Patients S6, S7 and S8 with homozygous stop gain *HYDIN* variants all showed almost static, extremely stiff cilia in airway epithelial cell samples, which is in line with previous studies of patients with *HYDIN* mutations. The patient S10 with heterozygous *HYDIN* variants had stiff cilia with some unsynchronized movement, which may be due to residual HYDIN in some of the axonemal central pair projections. All patients were reported to have low NO and recurrent sinusitis and otitis (Table 1). No situs inversus was reported for any of the patients as is expected for individuals carrying *HYDIN* mutations based on the lack of central pair structure in nodal cilia. Patient S8 with homozygous stop gain variants had developed bronchiectasis. We conclude that the symptoms reported for the patients can be explained by the identified *HYDIN* mutations and that this information can be used for genetic counselling and management of PCD in these patients.

## Discussion

Although genetic variants for PCD have been identified in over 60 genes across the world, the genetics of PCD is population specific in many cases; the majority of mutations causing PCD are found only in a single family. However, more widespread mutations have been identified within populations and across different ethnicities and geographical populations (Fassad, Patel et al. 2020, Hannah, Seifert et al. 2022, Rumman, Fassad et al. 2023, Xu, Feng et al. 2023). Thus, it is important to identify the population specific variants to improve diagnostics and management of the disease. Identification of genetic causes of a disease enables better prediction of the evolvement of symptoms in each patient and development of treatments including personalised medicines to improve their quality of life. There is tissue specificity and different severity of disease progression depending on the genes affected and types of mutations. The most severe PCD phenotypes arise from genes affecting multiciliogenesis (*CCNO, MCIDAS*) due to either complete lack or an inefficiently low number of motile cilia in the airways. Although, these genes do not affect sperm tail formation, they may affect fertility via lack of efferent duct cilia (Terre, Lewis et al. 2019, Hoque, Kim et al. 2022). Genotype-phenotype relationships have also been identified in large-scale topological data analysis (Berger, Cancian and Meyer 2012). Disease severity also appears to be significantly worse in patients with mutations in the molecular ruler genes *CCDC39* and *CCDC40* compared to most other structural gene mutations in PCD, or mutations introducing more subtle axonemal defects such as *DNAH11* mutations.

*HYDIN* mutations are relatively common in PCD patients, although clinical diagnosis of these patients has been hindered due to a lack of clear structural changes in the axoneme, lack of situs inversus and a pseudogene hindering the sequence-based diagnostics. Recently electron tomography and immunofluorescence microscopy have provided tools to confirm the structural defect in patients with *HYDIN* mutations and demonstrating the loss of HYDIN at the central pair complex through SPEF2 staining. PCD Detect has been developed based on image averaging and enhancement as a diagnostic tool kit to assess cases which are difficult to diagnose, but have a suggestive HSVA result, TEM result, or a variant of unknown significance (Shoemark, Pinto et al. 2020). In this study we utilized PCD Detect and structural studies to confirm the causality of compound heterozygous mutations together with immunofluorescence and RNA experiments.

In Finland genetic mutations in PCD patients have previously been identified only in the *DNAH11* and *CFAP300* genes (Schultz, Elenius et al. 2020, Schultz, Elenius et al. 2022). The *HYDIN* variants identified in this study contribute to the understanding of the genetic background of PCD in Finland and enable development of genetics diagnostics within the Finnish population. Confirmation of the causality of *HYDIN* variants is complicated by the presence of the pseudogene *HYDIN2*. Here we have utilized RNA sequencing and electron tomography to confirm the compound heterozygous mutations in *HYDIN*, which couldn’t be verified with Sanger sequencing of DNA or by conventional TEM. The homozygous stop gain variant c.1797C>G was found in three PCD patients and is predicted to truncate the protein product by 4522 aa, which likely causes degradation of the protein by nonsense mediated decay (NMD). The variant is not present in the GenomAD database and based on the results of this study we concluded that this is the disease-causing mutation in these patients. The compound heterozygous variants in patient S10 were validated with RNA expression and sequencing, immunohistochemistry and EM tomography and the results showed convincing evidence that PCD is caused by HYDIN defects in this patient as well. The c.13801delG (rs752405406) variant is enriched in the Finnish population with an allele frequency of 0.0005309, and is specific to the European population (0.000001695). The c.12899G>C (rs200169224) variant is more common in the Finnish population (0.003242), but can also be found in European, African and American populations (0.0002078-0.001935). Thus, these variants can be expected to be a more common cause of PCD in the Finnish population. Our study has shown that *HYDIN* variants explain 22% of the analysed cases (4/16) in the Finnish patient cohort. Other recurrent PCD-causing variants were previously reported in *CFAP300* (17%, three families) and *DNAH11* (11%, two families) (Schultz, Elenius et al. 2020, Schultz, Elenius et al. 2022). There is still work to be done to establish a more complete gene panel for genetic diagnostics in Finland, but several Finnish population enriched variants have been identified thus far and this information can be used for improving clinical diagnostics for PCD.

## Data Availability

All data produced in the present study are available upon reasonable request to the authors

## Acknowledgements

The authors would like to acknowledge the funding for this research from BBSRC grant BB/V011251/1. AIS was also funded by Marie Sklodowska-Curie individual fellowship No. 800556 from the European Union’s Horizon 2020 Research and Innovation Programme. HMM was supported by NIHR GOSH BRC, Ministry of Higher Education in Egypt and acknowledges support from the BEAT-PCD network (COST Action 1407 and European Respiratory Society (ERS) Clinical Research Collaboration). MRF was supported by a Wellcome Trust Collaborative Award in Science (210585/Z/18/Z).

## Author contributions

TB produced the electron tomography images and analysed the sample with PCD-detect, MRF performed the variant calling, RS collected patient samples, VE collected clinical data, patient samples and performed HSVA, JSYL performed immunofluorescence staining, GF performed Sanger sequencing, RR prepared electron microscopy samples, MAM extracted RNA, HMM supervised the study, AIS wrote the manuscript, conducted imaging, Sanger sequencing, variant filtering and bioinformatic analysis. All authors contributed corrections and improvements to the manuscript.

## Conflict of interest statement

The authors declare no conflict of interest.

## Notes

### Competing Interest Statement

The authors have declared no competing interest.

### Funding Statement

BBSRC grant BB/V011251/1, Marie Sklodowska-Curie individual fellowship No. 800556 from the European Unions Horizon 2020 Research and Innovation Programme, NIHR GOSH BRC, Ministry of Higher Education in Egypt, the BEAT-PCD network (COST Action 1407 and European Respiratory Society (ERS) Clinical Research Collaboration) and a Wellcome Trust Collaborative Award in Science (210585/Z/18/Z).

### Author Declarations

The study was ethically approved by the University of Turku Ethics Committee (ETMK 69 2017), London Bloomsbury Research Ethics Committee approved by the Health Research Authority (08/H0713/82), and the referring hospitals.

