## Supplemental table 1 for "*HYDIN* variants cause primary ciliary dyskinesia in the Finnish population"

Supplementary table 1. Primers used for Sanger sequencing and RT-PCR.

| **DNA** | **Forward** | **Reverse** | **length** | **Specificity** |
| --- | --- | --- | --- | --- |
| Hydin e14:c.C1797G | CTGCTGACACTGGACATGCTA | GACAGGTCTAGGACACACTCTATCA | 851 | *HYDIN1* |
| Hydin e76:c.G12899C | GGGGACAGGGACCTAGGATT | TGGAAATGGCACCCTGAGAC | 452 | *HYDIN1/HYDIN2* |
| Hydin e80:c.G13800C | GGGGACAGGGACCTAGGATT | TGGAAATGGCACCCTGAGAC | 452 | *HYDIN1/HYDIN2* |
| **RNA** |  |  |  |  |
| Hydin e76-79 | TTTCACCCATCGACAGCACT | CGGGGCAAAGATGACTTCCA | 713 | *HYDIN1* |
| Hydin_e79-80 | TGGACCCGTGGTGTATCAGA | AGACTCAGAGGACTGCCTCC | 249 | *HYDIN1* |
| Hydin_e79-81 | GGTGTATCAGACGCAAGCCA | AGATGGGGTGCAGATTCCAG | 377 | *HYDIN1* |
| Hydin_e85-86 | TTACACACGGCAGAGGACAG | GTGGCTGGGCTCGAATAAGA | 136 | *HYDIN1* |
| GAPDH | GTCATCCCTGAGCTGAACGG | AAGTGGTCGTTGAGGGCAAT | 263 | *GAPDH* |
